# An assessment of the vaccination of school-aged children in England

**DOI:** 10.1101/2022.02.04.22270361

**Authors:** Matt J. Keeling, Sam E. Moore

## Abstract

**Background:** Children and young persons are known to have a high number of close interactions, often within the school environment, which can facilitate rapid spread of infection; yet for SARS-CoV-2 it is the elderly and vulnerable that suffer the greatest health burden. Vaccination, initially targeting the elderly and vulnerable but later expanded to the entire adult population, has been transformative in the control of SARS-CoV-2 in England. However, early concerns over adverse events and the lower risk associated with infection in younger individuals means that the expansion of the vaccine programme to those under 18 year of age needs to be rigorously and quantitatively assessed.

**Methods:** Here, using a bespoke mathematical model matched to case and hospital data for England, we consider the potential impact of vaccinating 12-17 and 5-11 year olds. This analysis is reported from an early model (generated in June 2021) that formed part of the evidence base for the decisions in England, and a later model (from November 2021) that benefits from a richer understanding of vaccine efficacy, greater knowledge of the Delta variant wave and uses data on the rate of vaccine administration. For both models we consider the population wide impact of childhood vaccination as well as the specific impact on the age-group targeted for vaccination.

**Results:** Projections from June suggested that an expansion of the vaccine programme to those 12-17 years old could generate substantial reductions in infection, hospital admission and deaths in the entire population, depending on population behaviour following the relaxation of control measures. The benefits within the 12-17 year old cohort were less marked, saving between 656 and 1077 (95% prediction interval 281-2260) hospital admissions and between 22 and 38 (95% PI 9-91) deaths depending on assumed population behaviour. For the more recent model, the benefits within this age group are reduced, saving on average 631 (95% PI 304-1286) hospital admissions and 11 (95% PI 5-28) deaths for 80% vaccine uptake, while the benefits to the wider population represent a reduction of 8-10% in hospital admissions and deaths. The vaccination of 5-11 year olds is projected to have a far smaller impact, in part due to the later roll-out of vaccines to this age-group.

**Conclusions:** Vaccination of 12-17 year olds and 5-11 year olds is projected to generate a reduction in infection, hospital admission and deaths for both the age-groups involved and the population in general. For any decision involving childhood vaccination, these benefits needs to be balanced against potential adverse events from the vaccine, the operational constraints on delivery and the potential for diverting resources from other public health campaigns.

## Background

The SARS-CoV-2 pandemic has lead to severe healthcare burdens, often necessitating unprecedented non-pharmaceutical intervention measures[1**? ?**, 2]. From December 2020, the availability of highly effective vaccines (first the Pfizer-BioNTech, followed by the Oxford-AstraZeneca and Moderna vaccines) offered an alternative method for limiting infection and disease[3–5]. In England, this led to the largest ever vaccine campaign with around 100 million doses administered in the first twelve months. These vaccines were offered in a targeted manner, with healthcare workers, the vulnerable and the elderly being the first to be offered their initial dose[6].

One of the key characteristics of the SARS-CoV-2 pandemic has been the striking age-dependent nature of disease severity, with older (and clinically vulnerable) individuals suffering a far higher burden of hospital admissions and deaths [7–10]. Many countries initially closed schools as a response to this pandemic [11–14], but the role played by school-aged children and the strength of transmission within the school environment remain deeply contested with different conclusions being drawn from a range of modelling and data-driven analyses [15–31]. This dichotomy of views on the role of schools may be attributed to two conflicting factors. Firstly, all the available data highlights that younger individuals have a lower susceptibility to SARS-CoV-2 infection, a reduced risk of displaying symptoms and a greatly reduced risk of severe illness [7–9, 31]. However, it is also well established that the school environment is historically associated with a high transmission risk for many respiratory pathogens [32– 36], simply due to the large number of pupils and the high number of contacts involved [37–39]. The data from England suggests that (until the Omicron variant) there had not been the types of explosive outbreaks of SARS-CoV-2 within schools that are frequently associated with other respiratory diseases [40–43]; yet it is evident that transmission does occur within schools [44–46] and much of 2021 saw pronounced increases in school-aged infection in England [42, 47].

Here a complex age-structured model, matched to data in the seven NHS regions of England [48– 50], is used to examine the vaccination of 12-17 and 5-11 year olds. In particular, the number of infections, hospital admissions and deaths in the whole population and in the target age-groups are compared with and without vaccination. The model has been extensively used throughout the pandemic, particularly for questions about vaccination [50, 51] and to project the likely impact of relaxing control measures [52, 53]. This model formulation has also been used to consider the impact of school reopening on the population level incidence [15], although bespoke individual-scale models may be more appropriate when focusing on within-school dynamics rather than their feed-back on the community [24, 54, 55].

We consider the projected dynamics and the benefits from the perspective of a pre-Omicron assessment. The rapid spread of the Omicron variant has dramatically shifted the epidemiological landscape with far more infections expected in England during very early 2022. However, there are currently too many uncertainties surrounding fundamental parameters associated with this new variant to make robust quantitative projections.

The aim of this paper is not to argue for or against vaccination of particular age-groups, but to present a quantitative assessment of the epidemiological role that vaccination has played and is likely to play in the near future. As such the paper focuses on the positives of vaccination: the impact of vaccinating children and young people on the infection in the vaccinated age-group in particular and the population in general. There is no quantitative consideration or comparison of these benefits against potential risks or costs.

## Methods

Three sets of simulations are performed: one from 26th June 2021 which was used as part of the JCVI decision-making process for the vaccination of 12-17 year olds; one using data and information up to 6th November 2021 again looking at the benefits of vaccinating 12-17 year olds; and a final simulation based on 6th November data examining the benefit of vaccinating 5-11 year olds.

The model used throughout this paper is an age-structured formulation that has been developed throughout this pandemic [15, 48, 53] and has been continually matched to a range of epidemiological data [49]. The model is explained in more detail in the Supplementary Material, but a brief overview of the main characteristics are given here. The model is a deterministic age-structured model, partitioning the population into twenty-one 5-year age classes and partitioned spatially into the seven NHS regions of England. The model captures age-dependent mixing, susceptibility, likelihood of symptoms and likelihood of severe disease. It has been adapted throughout the course of the pandemic to include multiple variants [56] and vaccination[6, 50]. The November model also includes the waning of vaccine protection and infection-derived immunity as well as the deployment of booster vaccination. One of the principal drivers of epidemic behaviour is the mixing of individuals in the population, which can be modified by either as a result of imposed restrictions or voluntary behaviour change due to perceived risk. This level of precautionary behaviour is inferred as a slowly varying parameter by matching to recorded numbers of community test results, hospital admissions and hospital bed occupancy, ICU occupancy and deaths in each of the seven spatial regions.

The model is used to investigate the number of infections, hospital admissions and deaths using a weighted sum over a given time frame (we define *w*_*d*_ as the weighting applied on day *d*). Numbers with and without the vaccination of children are compared, considering both the total numbers across the entire population and numbers in the cohort being vaccinated. The results generated in June 2021 and presented to JCVI used a time window of 19th July 2021 until 31st December 2021 and weighted all time-points equally - with 19th July chosen as the start of Step 4 and the lifting of restrictions. However, it is arguably more appropriate to consider a longer time window (19th July 2021 until 31st December 2022) so that the longer impact of immunisation can be captured. We do this by having a constant weighting applied from 19th July to 31st December 2021, as previously, followed by weights that decrease linearly to zero by 31st December 2022. This gradual decline accounts for the greater uncertainty in longer-term projections and has the advantage over a discrete step-change that the results are relatively insensitive to the timing of future waves of infection. In the earlier step-change formulation a wave of infection in December 2021 would be included in the sum, but one in January 2022 would not; the use of a longer window with a gradual decline in the weighting overcomes many of these problems. The use of such a weighted time window draws into sharp focus the need to specify the total time frame over which any vaccine programme is evaluated; we quantify this by calculating the programme weighting, *T*_*V*_, which is the total weighted time over which the vaccination of the target cohort operates:

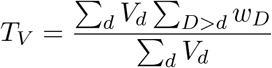

where *V*_*d*_ is the number of individuals in the cohort vaccinated on day *d*.

### June Projections

The model from the 26th June, was based on the model framework used for the 6th July Roadmap [53]. Vaccination of 12-17 year olds was included as part of the national vaccine roll-out, which was assumed to administer 2 million doses a week from late July onwards, such that 12-17 years would be offered vaccination once older groups had been offered theirs. Throughout it was also assumed that 80% of this age-group would take the vaccine. Given the need to first vaccinate those over 18 years old and to offer second doses at the correct time, the vast majority of first doses in the 12-17 age group take place between early August and late September 2021. It was assumed that second doses are given no sooner that 8 weeks after the first dose, with precise timing depending on the vaccine schedule leading to the majority of second doses in 12-17 year olds being administered between early October and late November 2021 (Figure 1).

**Fig. 1:**
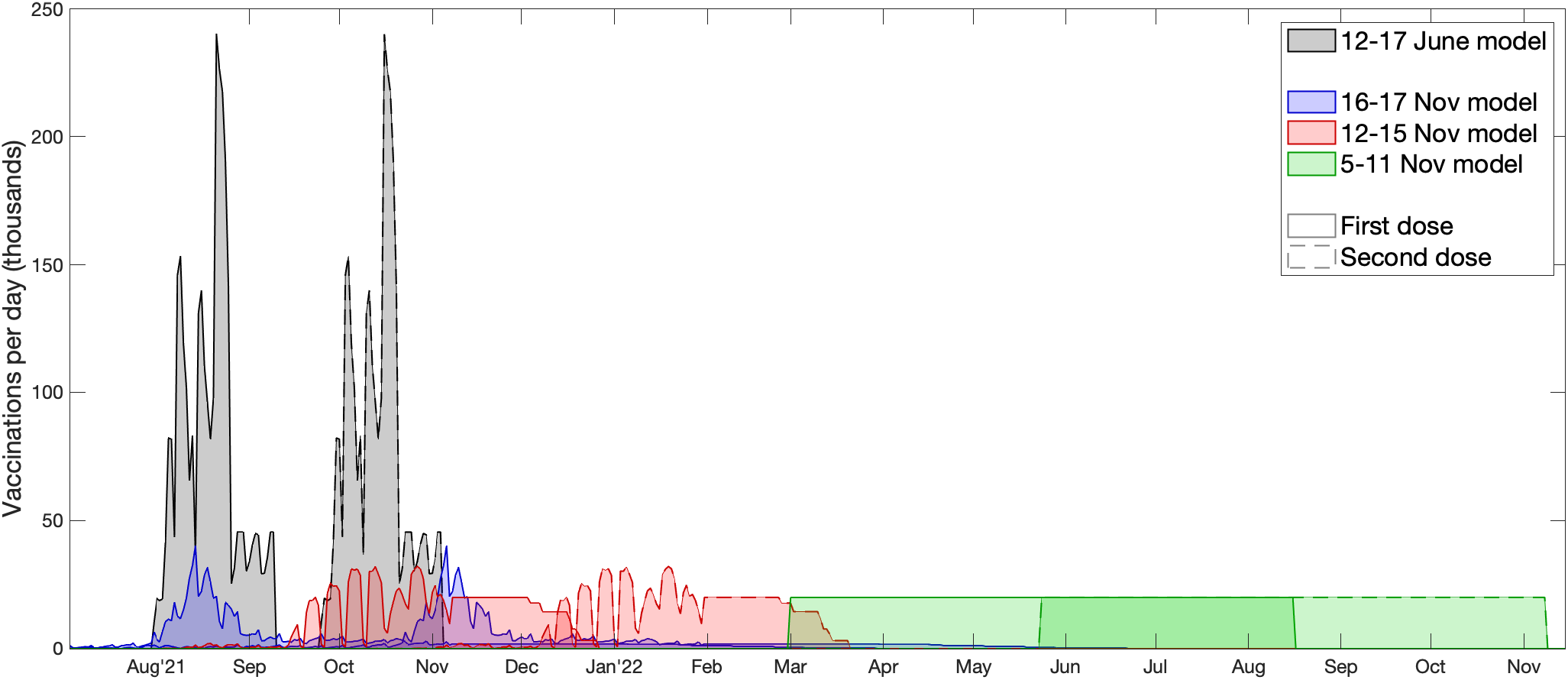
Timing of vaccine delivery in the two models (June: black, November: blue, red, green) by target age-group. Solid outlines correspond to first doses, while dashed outlines correspond to second doses. For the November model, results are shown for the default assumption of 140,000 vaccines per week through the school delivery programme, and a 70% uptake.

In late June 2021 the vaccine efficacy against the newly emerged Delta variant remained highly uncertain, with the best estimates suggesting limited impact from the first dose but markedly increased protection from the second dose. Although 12-17 year olds were largely given the Pfizer vaccine, Table 1 presents vaccine efficacy values from the time for both AstraZeneca and Pfizer as the protection from AstraZeneca is important for the rest of the population.

**Table 1:** Estimated vaccine efficacy (from the June projections) against infection, symptomatic disease, hospital admission and death after one or two doses with either the AstraZeneca ChAdOx vaccine or an mRNA vaccine such as the Pfizer vaccine. Values were taken from PHE reports at the time.

| Protection against: | AstraZeneca |  | Pfizer |  |
| --- | --- | --- | --- | --- |
|  | Dose 1 | Dose 2 | Dose 1 | Dose 2 |
| Infection | 34% | 71% | 34% | 73% |
| Symptoms | 34% | 82% | 34% | 83% |
| Hospitalisation | 64% | 90% | 64% | 91% |
| Deaths | 60% | 96% | 60% | 96% |

These June simulations were performed before ‘Step 4’ (on 19th July 2021) and the complete relaxation of all restrictions on social mixing; therefore a range of future possibilities were explored from an abrupt return to pre-COVID social mixing to a gradual return over several months. This uncertainty in population behaviour determines the scale and shape of the Delta wave, with more rapid returns to pre-COVID mixing leading to higher but shorter waves of infection.

Simulation results are aggregated either across the short-term time window (19th July - 31st December 2021, with equal weighting on all days) or the longer-term window (with constant weighted from 19th July - 31st December 2021, and then a linearly declining weighting until 31st December 2022). For vaccinations that begin in early August, this means that the short-term window has a programme weighting of 134 days over which childhood vaccination can impact the dynamics, which extends to around 316 days for the longer-term window.

### November Projections

The more recent model uses data and parameters up to 6th November to perform similar analyses, this data includes updated vaccine efficacy estimates (Table 2) which provide greater protection especially after the first dose compared to the earlier estimates. However, by this point in time, data were also accumulating on the efficacy of vaccines over time, with a strong signal of waning protection. Given this observation, from September 2021 booster (third) doses were offered to individuals who had received their second dose of vaccine more than six months ago. Both waning and boosters are included in the updated model framework [51].

**Table 2:**
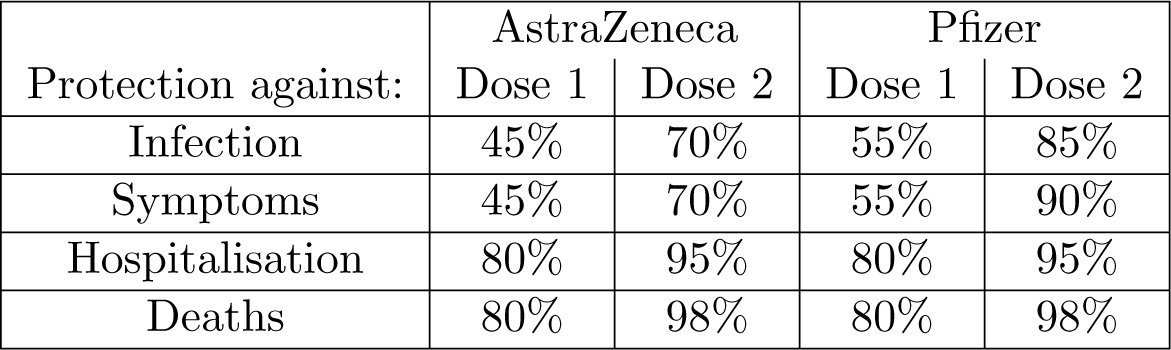
Estimated vaccine efficacy from the November models against infection, symptomatic disease, hospital admission and death after one or two doses with either the AstraZeneca ChAdOx vaccine or an mRNA vaccine such as the Pfizer vaccine. Values are taken from UKHSA reports [57, 58]

Given that the uncertainties in behaviour associated with the transition to Step 4 have been resolved, the November model focuses on a range of other unknowns including the speed of vaccine delivery and the uptake. By this point it was known that vaccination of 12-15 year olds was taking place in the school environment separating it from the logistical demands of vaccinating the rest of the population, while first doses had been offered to 16-17 year olds since August 2021. It is generally considered more equitable to vaccinate children though a school-based system, which also generally achieves better up-take than other methods. In these simulations the counterfactual scenario in which 12-17 year olds are not vaccinated is first considered; this is compared to projections that use the observed vaccinations until 6th November (by which time 70% of 16-17 year olds and 40% of 12-15 year olds have received one dose) and then continue vaccinating 12-15 year olds into the future until the required uptake (60%, 70% or 80%) is achieved (Figure 1). Sensitivity to the rate at which the vaccine roll-out continues from 6th November onwards is explored, vaccinating either 100,000, 140,000 or 180,000 12-15 year old pupils per week. For simplicity it is assumed that a second dose is given 12 weeks after the first (Figure 1).

For these November simulations only the longer-term window is used (with constant weighting from 19th July - 31st December 2021, and then a linearly declining weighting until 31st December 2022). Given that vaccination of 12-15 year olds began on 20th September 2021 and continued throughout 2021, this means that childhood vaccination has a programme weighting of 267 days.

This more recent model is also used to examine the vaccination of 5-11 year olds, assuming that this begins in March 2022, once second doses of 12-15 years is largely complete – logistical constraints make it unlikely that school-based programmes for 12-15 year olds and 5-11 year olds could operate in parallel. Vaccination of this age-group is assumed to be through the school environment and a range of vaccine uptake levels (60%, 70% and 80%) and a range of deployment speeds (slow - 100,000 per week, default - 140,000 per week or fast - 180,000 per week) are considered as part of the sensitivity analysis (Figure 1). Given that there are approximately 87% more 5-11 year olds than 12-15 year olds in England, vaccination of this cohort is a far lengthier process. The longer-term window is again used, which given that vaccination does not begin until March 2022, means that vaccination of 5-11 year olds has a programme weighting of only 81 days.

## Results

We begin by comparing age-structured model output (from the June and November models) against the data in terms of the number of individuals admitted to hospital in ten different age-groups, from 0-5 to over 85 years old (Figure 2). The June results (which are an average over the seven Step 4 scenarios investigated [53], including vaccination of 12-17 year olds) highlight an underestimation of the epidemic scale, but an overestimation of young people being hospitalised - this is principally due to the inability to forecast population level mixing in response to the Step 4 relaxation step. For the November simulations, there is a much closer agreement between model results and the data. These later replicates were performed by fitting to the total hospital admissions (without age-structure) over this period, so have been matched to the historic changes in imposed restrictions and precautionary mixing that drive the dynamics. Nevertheless, this comparison acts as a relatively independent assessment of the model fit, as age-structured data is not used as part of the inference process.

**Fig. 2:**
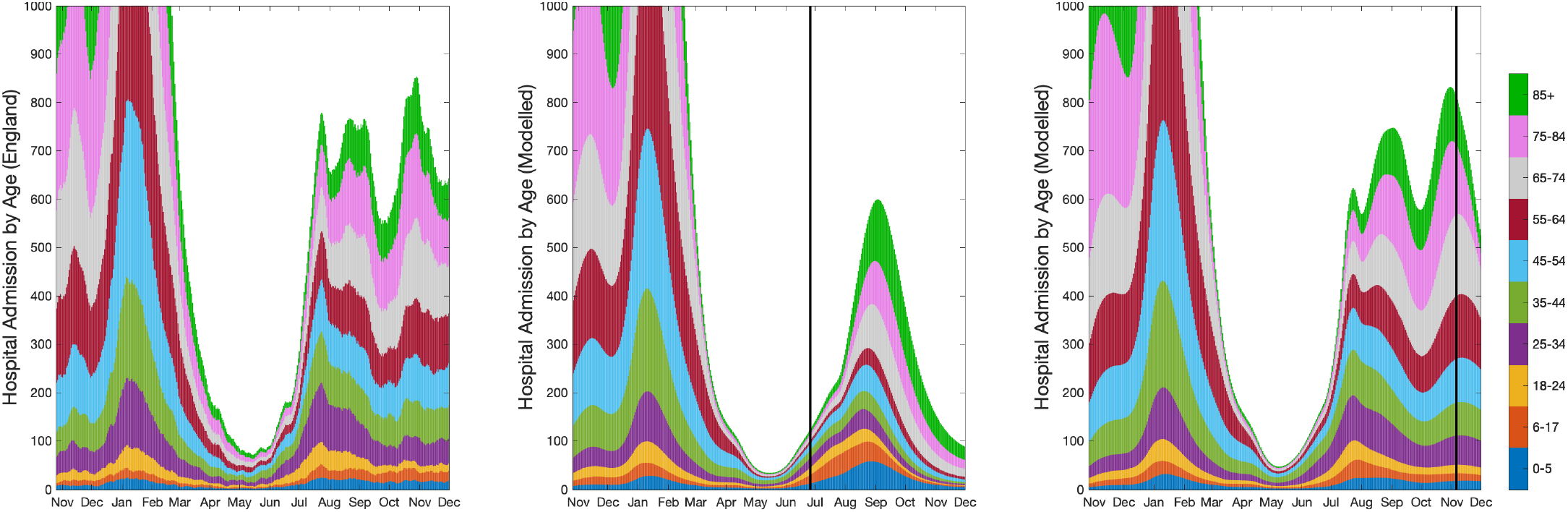
Comparison of data (left) and model results (June: centre, November: right) for the number of daily hospital admissions in ten age groups, from 0-5 years to 85 and above. The data on the left is a one-week moving average (using 3 days either side of the date) which removes any day-of-the-week effects and helps to smooth stochastic fluctuations. The model results are from the both June (average over seven Step 4 mixing scenarios) and November predictions and are computed using 5-year age-classes, and as such the first three age-groups shown are composites of multiple simulated age-classes: the 0-5 age group contains all projected hospital admissions from the 0-4 age-class and one fifth of all admissions from the 5-9 age-class. The vertical lines corresponds to the simulation dates for the two projections - 26th June and 6th November 2021.

### June Predictions

The projections generated in June 2021 had to make assumptions about the population mixing that would occur under Step 4 of the relaxation roadmap when all legal restrictions on social mixing were removed. A range of scenarios were investigated (bottom row, Figure 3), from an abrupt return to pre-COVID mixing (shown in purple and pink) to a far more gradual return (shown in green and cyan). Unsurprisingly, a rapid return to pre-COVID mixing leads to a single sharp peak in infection and hospital admissions, whereas a more gradual return often leads to a multi-peaked wave. These plots of the total number of hospital admissions show results without (solid line) and with (dashed line) the vaccination of 12-17 year olds.

**Fig. 3:**
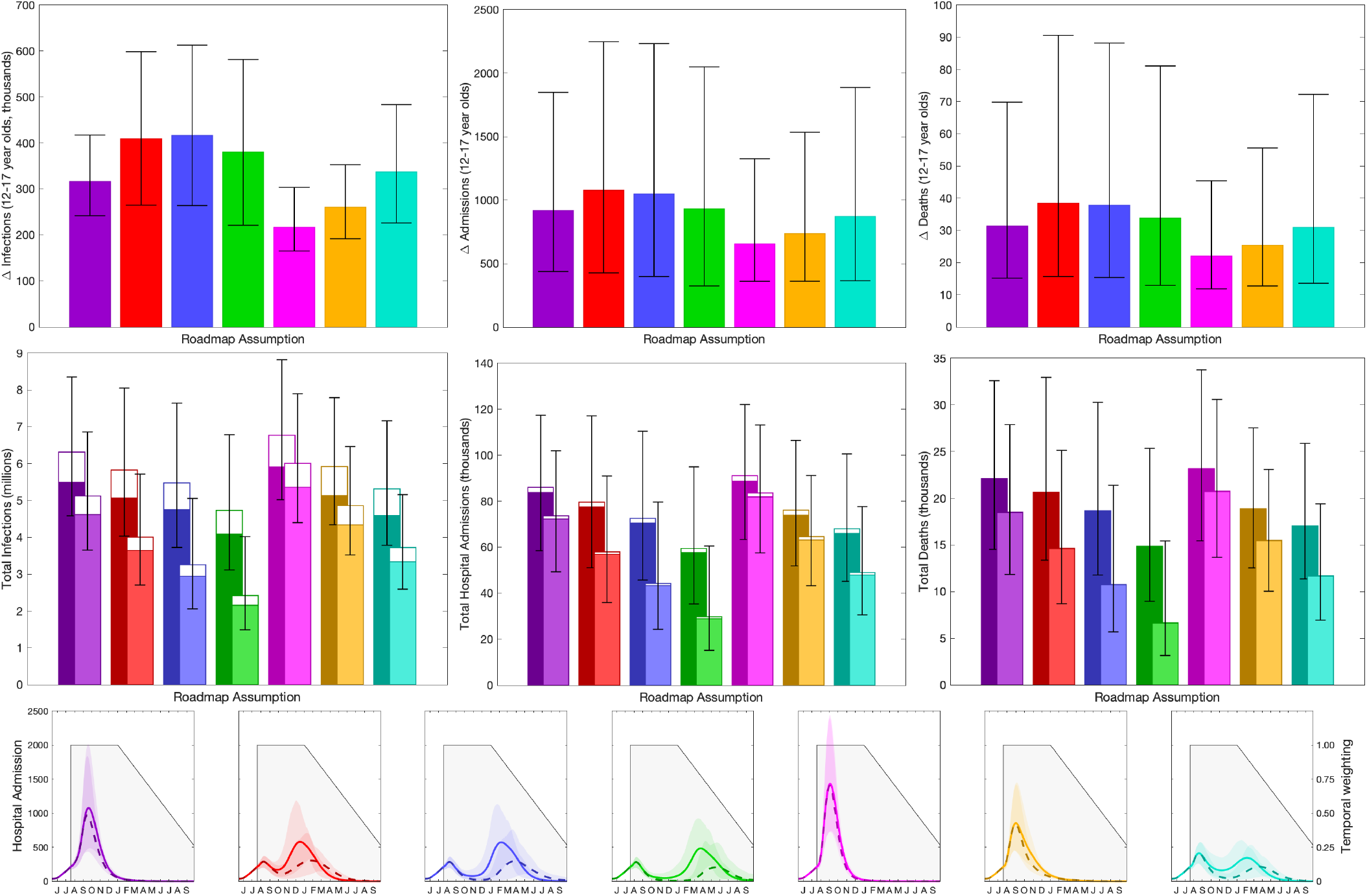
Impact of vaccination of 12-17 year olds in England, calculated in June 2021. Top row: reduction in infections, hospital admissions and deaths in 12-17 year olds due to vaccination in this age-group. Middle row: total number of infections, hospital admissions and deaths in the entire population (total bar) and 12-17 year olds (open bar). Lower row: number of projected hospital admissions over time (lines and ribbons), and the assumed time discounting (grey shading). In the top two rows bars are the mean value, error bars are the 95% prediction intervals, and different colours represent different assumptions about precautionary mixing in Step 4 [53]

The impact of childhood vaccination on the age-classes concerned (top row, Figure 3) leads to a reduction in the total number of infections (left), hospital admissions (centre) and deaths (right) that are projected to be accrued over the period 19th July 2021 to 31st December 2022, with the appropriate temporal weighting. Similar results but only over the period 19th July 2021 to 31st December 2021 are given in the Supplementary Material. The assumption about relaxation to pre-COVID behaviour (different colours in Figure 3) has a large effect on the dynamics and hence the impact of vaccines on the 12-17 year old age group. In general the results suggest that vaccination of 12-17 years reduces infection in the same age group by 200-400 thousand, reduces hospital admissions by around 600-1100, and reduces death by around 20-40 individuals, although the 95% prediction intervals on each of these are large showing the considerable uncertainty in the dynamics.

Broadening our scope to the entire population (middle row, Figure 3) shows the total number of infections, hospital admission and deaths over the time frame, both with (front lighter bars) and without (rear darker bars) vaccination of 12-17 year olds. The top bar that is unfilled corresponds to the numbers in the 12-17 age group, hence the difference between the two unfilled bars is the value shown in the top row. Error bars refer to the uncertainty in the total values. As expected, the assumptions about mixing after Step 4 on 19th July 2021 have a substantial impact on the total numbers predicted and the impact of vaccinating 12-17 year olds. Slow relaxation to pre-COVID mixing (e.g. green) leads to both lower totals and a greater impact of vaccination as the longer second (relaxation) wave generated when *R* just exceeds one is suppressed by vaccination. In contrast, for rapid relaxation (e.g. pink) there is insufficient time for the vaccination of 12-17 years to substantially reduce the infection dynamics. These graphs also highlight the lower severity of disease generally experienced by younger age groups; the fraction of each bar that is unfilled (representing those aged 12-17 years old) decreases substantially from infection, to hospital admission, to death.

### November Predictions, 12-17 year olds

Rerunning similar analysis in November, with greater understanding of vaccine efficacy against the Delta variant and hindsight for the social mixing after Step 4, leads to broadly similar patterns (Figure 4.)The lower panels again show examples of the total number of daily hospital admissions projected over time, together with the temporal weighting that is applied between 19th July 2021 and 31st December 2022. In these panels the black curve (and associated 95% prediction interval) refers to the counterfactual epidemic in the absence of childhood vaccination, while the blue, green and red curves show the impact of childhood vaccination with a slow, default and fast roll-out from November onward, respectively.

**Fig. 4:**
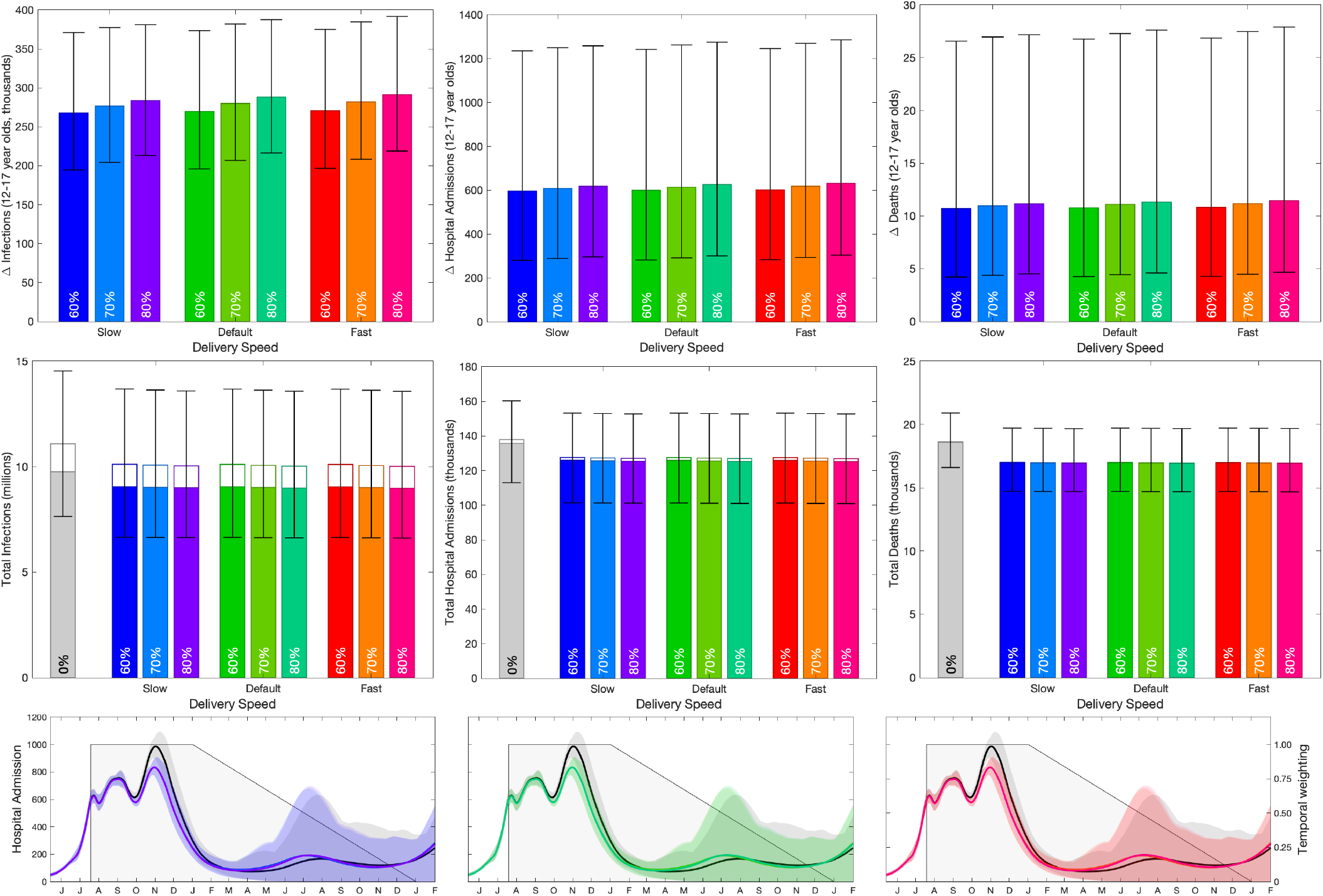
Impact of vaccination of 12-17 year olds in England, calculated in November 2021. Top row: reduction in infections, hospital admissions and deaths in 12-17 year olds due to vaccination in this age-group. Middle row: total number of infections, hospital admissions and deaths in the entire population (total bar) and 12-17 year olds (open bar). Lower row: number of projected hospital admissions over time (lines and ribbons; black for without childhood vaccination, colours corresponding to the bars above with 70% uptake and default speed), and the assumed time discounting (grey shading). In the top two rows bars are the mean value, error bars are the 95% prediction intervals, and different colours represent different assumptions about uptake in 12-15 year olds (no vaccination, 60%, 70% and 80%) and different assumptions about vaccine delivery speed (slow - 100,000 per week, default - 140,000 per week, fast - 180,000 per week). In 16-17 year olds, uptake was based on observations up to mid November except the counterfactual of no vaccination in which case all vaccination in 12-17 year olds were removed.

Focusing on the implications for vaccination of 12-17 year olds on this age-group (top row, Figure 4), delivery speed (blue, green and red) has relatively limited impact on the dynamics - in part because the vaccination of 12-17 year old up to 6th November 2021 is set by the observed schedule, which also restricts the timing of second doses. There is a more pronounced impact of vaccine uptake, with higher uptakes generating a greater impact. As seen with the earlier projections, there is a far greater reduction in infections compared to hospital admissions compared to deaths. Contrasting the June and November results, shows a lower benefit (approximately halved) in all three measures from the revised November model compared to the results from June. This is partly attributable to the larger wave of infection from July to December 2021 in the November simulations compared to some of the June scenarios; in terms of epidemic scale the November simulations most closely match the pink June scenario which has the lowest impact of vaccinating 12-17 year olds.

When considering the impact of vaccinating 12-17 year olds on the entire population (middle row, Figure 4), this again highlights that while infections in the 12-17 age group are relatively common (unfilled bar compared to the total height), hospital admissions and deaths are far less frequent in this age-group. These plots compare the counterfactual model without childhood vaccination (grey bar) with the vaccination of 12-17 year olds at different deployment speeds (after 6th November) and with different uptakes. Compared to the June results, the impact of childhood vaccination is diminished in the November simulations, with none of the later results generating the larger reductions that were projected for some of the Step 4 scenarios. This reduced impact of vaccination can be attributed to multiple factors: the difference in the pattern and scale of infection between July and December 2021, the higher vaccine efficacy assumptions in November such that the majority of the older population have greater protection and the slower roll-out schedule compared to earlier estimates.

### November Predictions, 5-11 year olds

Switching attention to the vaccination of younger children aged 5-11 years, it is assumed that their vaccination begins in March 2022, by which time the majority of older children (12-17) will have already been offered two doses. This later start date, coupled with the temporal weighting applied to all sums, means that there is less time (and less weight at each time point) for the vaccine to register an impact; the programme weighting has been reduced to 81 days. Therefore, while it makes sense to compare different scenarios for vaccination of 5-11 year olds, it is difficult to compare between the two childhood programs (12-17 year olds vs 5-11 year olds) due to the different time-frames involved - earlier vaccination will inevitably generate greater impacts.

Considering the impact of vaccinating 5-11 year olds on infection and disease in the 5-11 year old age-group (top panels, Figure 5) delivery speed (blue - slow, green - default, red - fast) has a pronounced impact as the entire delivery programme is now being simulated. The impact of vaccine uptake is most pronounced when the roll-out is fast, as this achieves the maximum coverage in the shortest amount of time, before these individuals become infected. For all these results, the lower bound of the 95% prediction intervals is close to zero, showing that in 2.5% of the projections the vaccination of 5-11 year olds is ineffective as there is predicted to be a substantial wave of infection in this age-group before vaccine delivery starts.

**Fig. 5:**
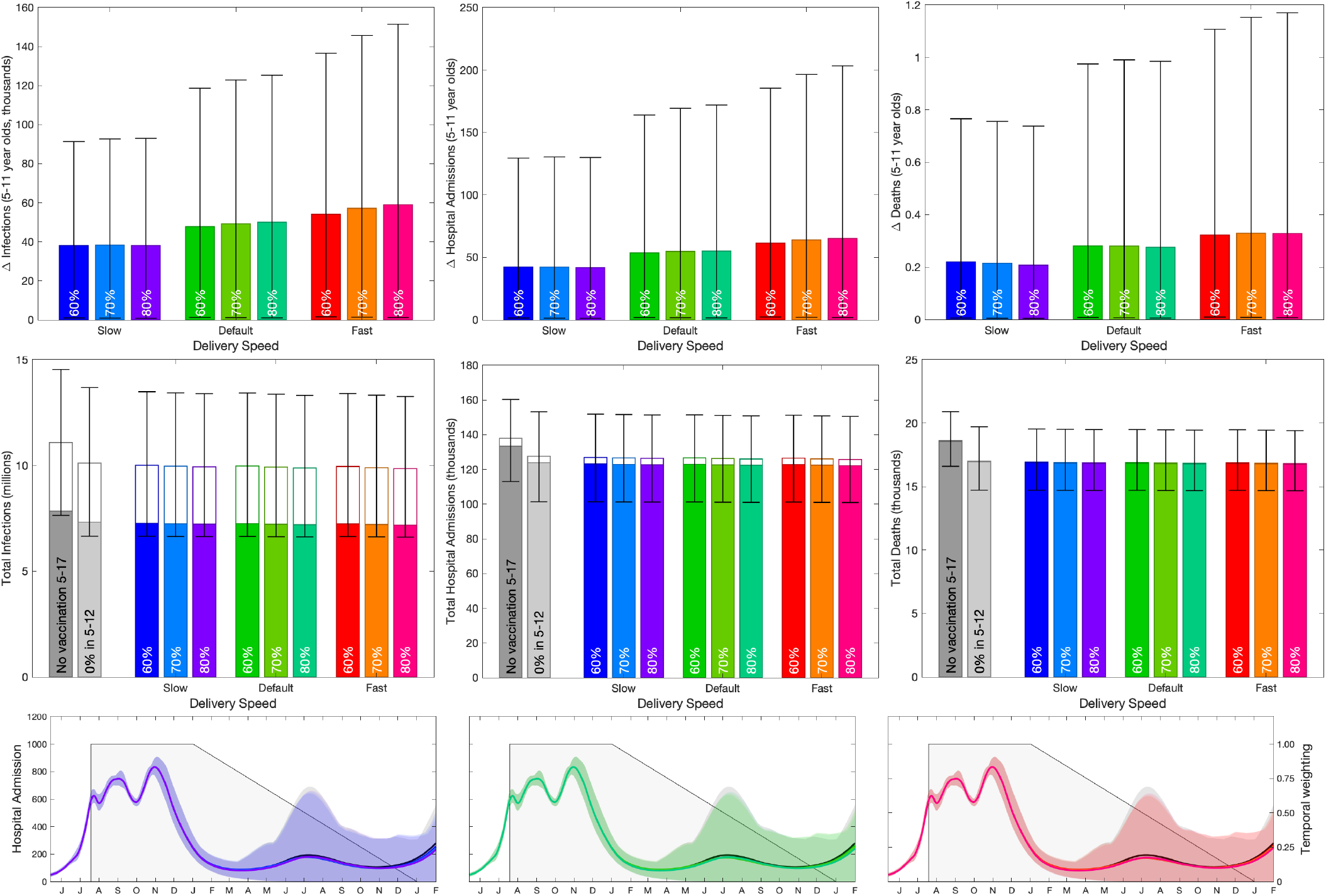
Impact of vaccination of 5-11 year olds in England, calculated in November 2021. Top row: reduction in infections, hospital admissions and deaths in 5-11 year olds due to vaccination in this age-group. Middle row: total number of infections, hospital admissions and deaths in the entire population (total bar) and 5-17 year olds (open bar). Lower row: number of projected hospital admissions over time (lines and ribbons; black for vaccinating those aged 12 and above but not 5-11 year olds, colours corresponding to the bars in the above rows), and the assumed time discounting (grey shading). In the upper two rows bars are the mean value, error bars are the 95% prediction intervals, and different colours represent different assumption about uptake in 5-17 year olds (no vaccination, 60%, 70% and 80%) and different assumptions about vaccine delivery speed (slow - 100,000 per week, default - 140,000 per week, fast - 180,000 per week). In the lower row, the counterfactual of no vaccination in all 5-17 year olds (dark grey) and the vaccination of 12-17 but not 5-11 (light grey) are both shown.

Again considering the impact on the entire population of vaccinating this youngest age-group (bottom row, Figure 5) highlights that vaccination of 5-11 year olds has a minimal effect on the rest of population, partly due to the late start to the vaccine campaign such that older individuals are already protected by immunization or infection.

## Discussion

The SARS-CoV-2 immunisation programme for children and young persons in England has evolved in an iterative manner, which is reflected in the timing of vaccinations in the later model. From late July (JCVI advice published 19th July 2021 [59]) those aged 12-17 with specific underlying health conditions that put them at risk of serious COVID-19 were offered the vaccine. This was extended in August (JCVI advice published 4th August 2021 [60]) to a first dose to all 16-17 year olds, a second dose 12 weeks later (JCVI advice published 15th November 2021 [61]) and a booster dose after 3 months (JCVI advice published 22nd December 2021 [62]). Those aged 12-15 (and not in a high-risk category) were offered their first doses through schools from late September (JCVI advice published 3rd September 2021 [63]), with a second dose 12 weeks later (JCVI advice published 29th November 2021 [64]). Finally, at-risk 5-11 year olds with specific underlying health conditions that put them at risk of serious COVID-19 were offered the vaccine from late December 2021 (JCVI advice published 22nd December 2021 [62]).

As can be seen from the decisions for England, there are considerable advantages in (initially) targeting vaccination to the most vulnerable[65, 66] - applying equally well to both adults and children. Available data suggests that clinically vulnerable children comprise around half of hospital admissions and three quarters of deaths in this age-group ([10, 67]), so protecting these individuals first with early targeted vaccination provides the greatest immediate impact. Due to a lack of available data, vulnerable groups have not been explicitly captured within our model; instead the risks (as captured by the infection:hospital ratio or infection:mortality ratio) are an average over all individuals within a five-year age-group. As such our results pertain to the advantages of vaccinating entire age cohorts; once the vulnerable have been offered the vaccine, the benefit of vaccinating the non-vulnerable is likely to be significantly reduced. Calculating the size of this reduction is not simply a matter of scaling down hospital admissions and deaths, but also needs to account for the indirect impact of vaccination (protection due to the reduction of infection) in this target age-group.

This paper has focused on the epidemiological impacts of vaccinating 5-17 year olds; calculating the reduction in infection, hospital admissions and deaths over a given time period. Using the November projections, we would estimate a reduction of approximately 250 thousand infections, 600 hospital admissions and 10 deaths in the 12-17 year old age-group over the time-period studied, if at least 60% of this cohort were vaccinated; and an additional reduction of 40-60 thousand infections and 40-60 hospital admissions in the 5-11 year old age-group when vaccinating this younger age. In part the lower numbers from the vaccination of 5-11 year olds comes from the lower programme weighting: 81 days compared to 267 days - due to the later impacts of the programme.

When considering the implementation of any new vaccine program it is important to consider the potential risks and costs as well as simply the expected benefits. In England, it is usually common to formulate a cost-benefit analysis in terms of financial costs of the vaccine against the benefit in terms of QALYs gained[68, 69] - although this approach has not been used when considering vaccination against SARS-CoV-2. In addition, there is now evidence that the general public expect the benefits of immunisation to substantially outweigh any health risk associated with adverse events of vaccination[70]. In this context, there were considerable early concerns about the risks of myocarditis and pericarditis following vaccination in younger individuals[71–74], and the potential for such adverse effects to disrupt the public’s confidence in the entire vaccination programme. However, there are clearly benefits associated with vaccination of 5-17 year olds; while the saving in terms of hospital admission and deaths is moderate compared to the total numbers observed so far, the benefits to the vaccine cohorts especially in terms of minimising infection and associated risks or minimising educational losses is clear.

There are several factors that could be introduced into the modelling framework to increase its realism. As mentioned above, being able to separate the younger population into vulnerable and non-vulnerable individuals would mean that the models would more closely replicate the pattern of vaccine roll-out deployed in England. While the model generates the number of infections and hospital admissions due to COVID-19, it does not attempt to quantify long-COVID - in part due to a lack of clinical definition and hence a lack of national-scale data[75, 76]. For some scenarios, the number of individuals involved is relatively small (e.g. the reduction in deaths in 5-11 year olds due to vaccinating 5-11 year olds) in which case it may be beneficial to adopt a stochastic approach such that the numbers predicted are all integers. Finally, the models do not include the Omicron variant; data on this new variant are still relatively sparse prohibiting the ability to make robust long-term projections. A substantial Omicron wave in early 2022 would likely increase the benefits of vaccinating 12-17 year olds as they would have protection against this increase in infection; however if would likely reduce the benefits of vaccinating 5-11 year olds as they are unlikely to be vaccination before any peak of the Omicron wave. The lower vaccine efficacy and lower severity reported for the Omicron variant could also reduce the benefits of childhood vaccination - more data is needed before detailed simulations can be performed.

## Conclusions

We have shown that vaccination of 12-17 year olds, and to a lesser extent the vaccination of 5-11 year olds, reduces infection, hospital admissions and deaths. These reductions were projected to be higher when modelling the dynamics in June compared to November 2021, partly due to an assumption of more rapid vaccine delivery and partly due to the unexpected scale of the third wave from July to December 2021. The projections from November 2021 suggest the vaccination of 12-17 year olds will generate a saving of around 600 (PI 280-1280) hospital admissions and a reduction of around 280,000 (PI 200,000-380,000) infections in this age group; while without vaccination we project 2240 (PI 1280-4000) hospital admissions and 1.3 million (PI 1.0-1.6 million) infections. The decision whether to offer vaccination to these younger individuals must balance the benefits to both the vaccinated age-group and the population in general against the risks of adverse events and the potential disruption to a range of other public health activities.

## Data Availability

Data from the CHESS and SARI databases were supplied after anonymisation under strict data protection protocols agreed between the University of Warwick and Public Health England. The ethics of the use of these data for these purposes was agreed by Public Health England with the Governments SPI-M(O) / SAGE committees.
Data on cases were obtained from the COVID-19 Hospitalisation in England Surveillance System (CHESS) data set that collects detailed data on patients infected with COVID-19. Data on COVID-19 deaths were obtained from Public Health England. These data contain confidential information, with public data deposition non-permissible for socioeconomic reasons. The CHESS data resides with the National Health Service (www.nhs.gov.uk) whilst the death data are available from Public Health England (www.phe.gov.uk). The ethics of the use of these data for these purposes was agreed by Public Health England with the Governments SPI-M(O) / SAGE committees. More aggregate data is freely available from the UK Coronavirus dashboard: https://coronavirus.data.gov.uk/

## Ethical considerations

Data from the CHESS and SARI databases were supplied after anonymisation under strict data protection protocols agreed between the University of Warwick and Public Health England. The ethics of the use of these data for these purposes was agreed by Public Health England with the Government’s SPI-M(O) / SAGE committees.

## Consent for publication

Not Applicable.

## Availability of data and materials

Data on cases were obtained from the COVID-19 Hospitalisation in England Surveillance System (CHESS) data set that collects detailed data on patients infected with COVID-19. Data on COVID-19 deaths were obtained from Public Health England. These data contain confidential information, with public data deposition non-permissible for socioeconomic reasons. The CHESS data resides with the National Health Service (www.nhs.gov.uk) whilst the death data are available from Public Health England (www.phe.gov.uk). The ethics of the use of these data for these purposes was agreed by Public Health England with the Governments SPI-M(O) / SAGE committees. More aggregate data is freely available from the UK Coronavirus dashboard: https://coronavirus.data.gov.uk/

## Patient and public involvement

This was an urgent public health research study in response to a Public Health Emergency of International Concern. Patients or the public were not involved in the design, conduct, or reporting of this rapid response research.

## Author Information

MJK performed the analysis, interpreted the results and was the major contributor in writing the manuscript. SM developed the early simulation software and contributed to the overall conception and design of the work. All authors read and approved the final manuscript.

## Additional Files

### Additional file 1 — June Projections with shorter time window

Figure showing the original assessment of vaccinating 12-17 year olds, shared with JCVI in June 2020, with used a much shorter and more abrupt time window - weighting all values between 19th July and 31st December 2021 equally.

### Additional file 2 — Model Formulation

Detail of the underlying mathematical framework that defines the transmission model. We break the model desciption into multiple sections that combine to generate a picture of SARS-CoV-2 transmission in the UK.

