## Additional Files 1 and 2 for "An assessment of the vaccination of school-aged children in England"

### Additional File 1: June Projections with shorter time window

In the original assessment of vaccinating 12-17 year olds, a much shorter and more abrupt time window was used - weighting all values between 19th July and 31st December 2021 equally (S1). Results for the mean number of infections (top left) and hospital admissions (top centre) saved in the 12-17 age-group and the mean number of hospital admissions (middle left) across the entire population were shared with JCVI.

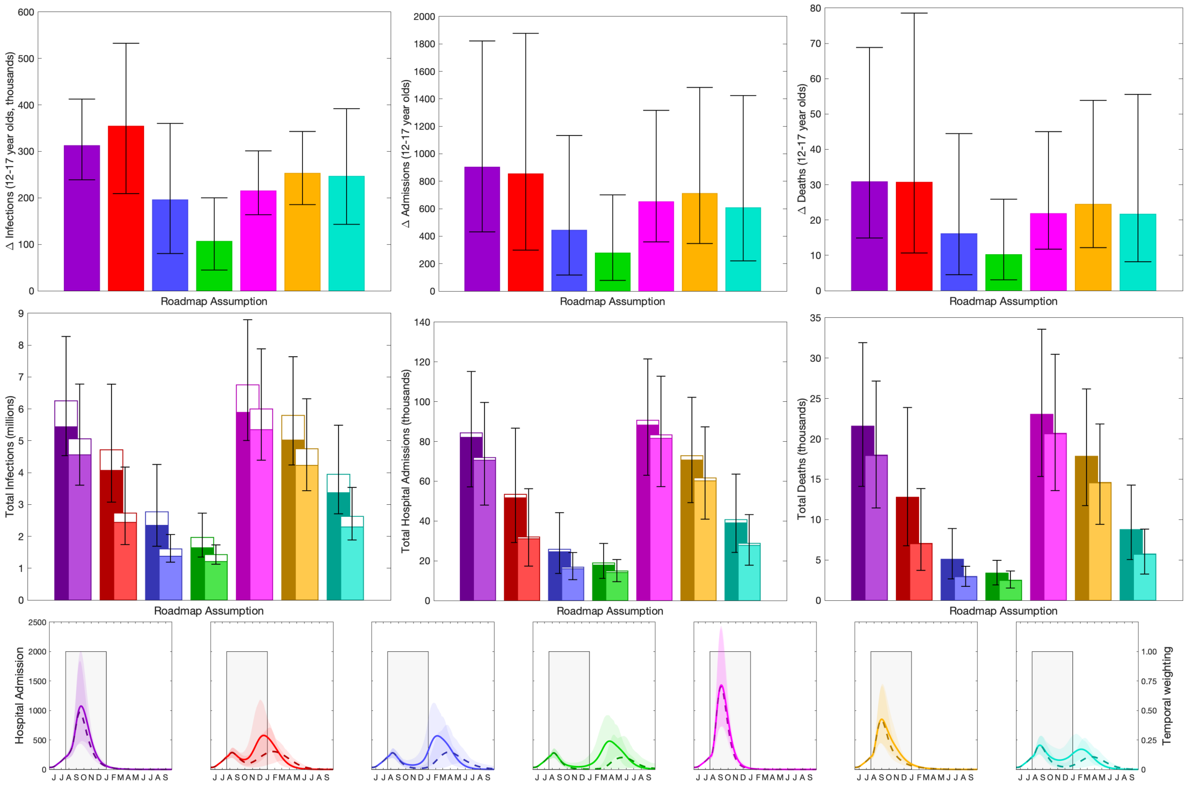


**Fig. S1: Impact of vaccination of 12-17 year olds in England, calculated in June 2021.** Top row: reduction in infections, hospital admissions and deaths in 12-17 year olds due to vaccination in this age-group. Middle row: total number of infections, hospital admissions and deaths in the entire population (total bar) and 12-17 year olds (open bar). Lower row: number of projected hospital admissions over time (lines and ribbons), and the assumed time discounting (grey shading). In the top two rows bars are the mean value, error bars are the 95% prediction intervals, and different colours represent different assumptions about precautionary mixing in Step 4 [53]. This is the equivalent of Figure 3 in the main text, but with quantities calculated over a shorter time-window.

### Additional File 2: Model Formulation

Here we detail the underlying mathematical framework that defines the transmission model. We break the model description into multiple sections that combine to generate a picture of SARS-CoV-2 transmission in the UK. This model structure has been detailed in previous publications [6, 15, 48, 50] but we review the details here for completeness.

#### Infection modelling

As is common to most epidemiological modelling we stratify the population into multiple disjoint compartments and capture the flow of the population between compartments in terms of ordinary differential equations. At the heart of the model is a modified SEIR equation, where individuals may be susceptible ($S$), exposed ($E$), infectious with symptoms ($I$), infectious and either asymptomatic or with very mild symptoms ($A$) or recovered ($R$). Both symptomatic and asymptomatic individuals are able to transmit infection, but asymptomatic infections do so at a reduced rate given by $\tau$. Hence, the force of infection is proportional to $I + \tau A$. To some extent, the separation into symptomatic ($I$) and asymptomatic ($A$) states within the model is somewhat artificial as there are a wide spectrum of symptom severities that can be experienced, with the classification of symptoms changing over time. Our classification reflects early case detection, when only relatively severe symptoms were recognised.

To obtain a better match to the infection time scales, we model the exposed class as a 3-stage process - this provides a better match to the time from infection to becoming infectious, such that in a stochastic formulation the distribution of the latent period would be an Erlang distribution.

$$
\begin{aligned}
\frac{dS}{dt} &= -\lambda S \qquad\qquad \text{where } \lambda \propto (I + \tau A) \\
\frac{dE_1}{dt} &= \lambda S - 3\alpha E_1 \\
\frac{dE_2}{dt} &= 3\alpha E_1 - 3\alpha E_2 \\
\frac{dE_3}{dt} &= 3\alpha E_2 - 3\alpha E_3 \\
\frac{dI}{dt} &= 3d\alpha E_3 - \gamma I \\
\frac{dA}{dt} &= 3(1-d)\alpha E_3 - \gamma A \\
\frac{dR}{dt} &= \gamma (I + A)
\end{aligned}
$$

where $\alpha^{-1}$, and $\gamma^{-1}$ are the mean latent and infectious periods, while $d$ is the proportion of infections that develop symptoms.

#### Age Structure and Transmission Structure

The simple model structure is expanded to twenty-one 5-year age-groups (0-4, 5-9, .... ,95-99, 100+). Age has three major impacts on the epidemiological dynamics, with each element parameterised from the available data:
Older individuals have a higher susceptibility to SARS-CoV-2 infection (captured by the parameter $\sigma$).
Older individuals have a higher risk of developing symptoms, and therefore have a greater rate of

transmission per contact.
Older individuals have a higher risk of more severe consequences of infection including hospital admission and death.

The age-groups interact through four who-acquired-infection-from-whom transmission matrices, which capture the epidemiological relevant mixing in four settings: household ($\beta_H$), school ($\beta_S$), workplace ($\beta_W$) and other ($\beta_O$). We took these matrices from Prem et al. [38] to allow easy translation to other geographic settings, although other sources could be used.

One of the main modifiers of mixing and therefore transmission is the level of precautionary behaviour, $\phi$ (see Figure 2 of the main text). This scaling parameter changes the who-acquired- infection-from-whom transmission matrices in each transmission setting, such that when $\phi = 1$ mixing in workplaces and other settings take their lowest value, whereas when $\phi = 0$ the mixing returns to pre-pandemic levels. Mixing within the school setting follows the prescribed opening and closing of schools.

$$\begin{aligned}
\frac{dS_a}{dt} &= -\lambda_a S_a \qquad\qquad \text{where } \lambda_a = \sigma_a \sum_b \beta_{ab}(\phi)(I_b + \tau_b A_b) \\
\frac{dE_{a1}}{dt} &= \lambda_a S_a - 3\alpha E_{a1} \\
\frac{dE_{a2}}{dt} &= 3\alpha E_{a1} - 3\alpha E_{a2} \\
\frac{dE_{a3}}{dt} &= 3\alpha E_{a2} - 3\alpha E_{a3} \\
\frac{dI_a}{dt} &= 3d_a\alpha E_{a3} - \gamma I_a \\
\frac{dA_a}{dt} &= 3(1-d_a)\alpha E_{a3} - \gamma A_a \\
\frac{dR_a}{dt} &= \gamma(I_a + A_a)
\end{aligned}$$

For simplicity of notation, we write the sum of the four age-structured mixing matrices as $\beta_\phi$.

To ensure that we can replicate the long-term dynamics of infection we allow the population to age. The aging process occurs annually (corresponding to the new school year in September) in which approximately one fifth of each age-group moves to the next oldest age cohort — small changes to the proportion moving between age-groups are made to keep the population size within each age- group constant.

#### Capturing Quarantining

One of the key characteristics of the COVID-19 pandemic in the UK has been the use of self-isolation and household quarantining to reduce transmission. We approximate this process by distinguishing between first infections (caused by infection related to any non-household mixing) and subsequent household infections (caused by infection due to household mixing). The first symptomatic case within a household (which might not be the first infection) has a probability ($H$) of leading to household quarantining; this curtails the non-household mixing of the individual and all subsequent infections generated by this individual.

In our notation, we let superscripts denote the first infection in a household ($F$), a subsequent infection from a symptomatic household member ($SI$) and a subsequent infection from an asymptomatic household member ($SA$); the first detected case in a household who is quarantined ($QF$) and all their subsequent household infections ($QS$). For a simple SEIR model (ignoring multiple E categories and age-structure) our extension would give:

$$\begin{aligned}
\frac{dS}{dt} &= -(\lambda^F + \lambda^{SI} + \lambda^{SA} + \lambda^Q)S \\
\frac{dE_1^F}{dt} &= \lambda^F S - 3\alpha E_1^F \\
\frac{dE_1^{SI}}{dt} &= \lambda^{SI} S - 3\alpha E_1^{SI} \\
\frac{dE_1^{SA}}{dt} &= \lambda^{SA} S - 3\alpha E_1^{SA} \\
\frac{dE_1^Q}{dt} &= \lambda^Q S - 3\alpha E_1^Q \\
\frac{dE_2^*}{dt} &= 3\alpha E_1^* - 3\alpha E_2^* \\
\frac{dE_3^*}{dt} &= 3\alpha E_2^* - 3\alpha E_3^* \\
\frac{dI^F}{dt} &= 3d(1-H)\alpha E_3^F - \gamma I^F \\
\frac{dI^{SI}}{dt} &= 3d\alpha E_3^{SI} - \gamma I^{SI} \\
\frac{dI^{SA}}{dt} &= 3d(1-H)\alpha E_3^{SA} - \gamma I^{SA} \\
\frac{dI^{QF}}{dt} &= 3dH\alpha E_3^F - \gamma I^{QF} \\
\frac{dI^{QS}}{dt} &= 3d\alpha E_3^Q + 3dH\alpha E_3^{SA} - \gamma I^{QS} \\
\frac{dA^X}{dt} &= 3(1-d)\alpha E_3^X - \gamma A^X \qquad \text{where } X \in \{F, SI, SA, Q\} \\
\frac{dR}{dt} &= \gamma \sum_X (I^X + A^X)
\end{aligned}$$

$$\begin{aligned}
\text{where } \lambda^F &= (\beta^S + \beta^W + \beta^O)(I^F + I^{SI} + I^{SA} + \tau A^F + \tau I^{SI} + \tau A^{SA}) \\
\lambda^{SI} &= \beta^H I^F \\
\lambda^{SA} &= \beta^H A^F \\
\lambda^Q &= \beta^H A^{QF}
\end{aligned}$$

This formation has been shown to be able to reduce R below one even when there is strong within household transmission, as infection from quarantined individuals cannot escape the household [19].

#### Spatial Modelling

Within England the model operates at the scale of NHS regions (East of England, London, Midlands, North East, North West, South East and South West). For simplicity and speed of simulation we assume that each of these regions acts independently and in isolation - we do not model the movement of people or infection across borders. In addition, the majority of parameters are regionally specific, reflecting different demographics, deprivation and social structures within each region. However, we include a hyper-prior on the shared parameters such that the behaviour of each region helps inform the value in others.

#### 0.1 Variant Modelling

The model also captures the three main variants that have been responsible for most infections in England: the wildtype virus (encapsulating all pre-Alpha variants), the Alpha variant and the Delta variant. Each of these requires a replication of the infectious states for each variant type modelled. We assume that infection with each variant confers immunity to all variants, such that there is indirect competition for susceptible individuals. This competition is driven by the transmission advantage of each variant which is estimated by matching to the proportion of positive community PCR tests (Pillar 2 test) that are positive for the S-gene. The TaqPath system that is used for the majority of PCR tests in England is unable to detect the S-gene in Alpha variants, due to mutations in the S-gene. The switch from S-gene positive to S-gene negative and back to S-gene positive corresponds with the dominance of wildtype, Alpha and Delta variants. We infer the transmissibility of Alpha and Delta variants to be 52% (CI 35-71%) and 156% (CI 117-210%) greater than wildtype, respectively.

#### Vaccination Modelling

We capture vaccination using a leaky approach, although non-leaky (all-or-nothing) models produce extremely similar results over the time-scales considered. The model replicates the action of:

- first and second doses of vaccine, at rates $v_1$ and $v_2$ respectively that move susceptible individuals through to vaccinated states ($VS_1$ and $VS_2$) but have no impact on infected or recovered individuals;
- waning vaccine efficacy at rates $\omega_1$ and $\omega_2$, giving a two-step process from fully vaccinated to waned efficacy (in the equation below, for simplicity we assume everyone who gets a first dose of vaccine also gets a second, so that waning from state $S_1$ is unnecessary);
- waning immunity at rates $\Omega_1$ and $\Omega_2$ which are assumed to be slower than the waning of vaccine efficacy.

In the June model waning was not included, hence $\omega_1 = \omega_2 = \Omega_1 = \Omega_2 = 0$. The model also needs to capture the total number of individuals who have been given a first or second dose of vaccine ($V_1$ or $V_2$ out of a total population size of $N$) to ensure that only individuals that have not been vaccinated are offered a first dose, and only individuals that have been vaccinated once are offered a second dose.

$$\frac{dS}{dt} = -v_1 \frac{S}{N - V_1}$$
$$\frac{dVS_1}{dt} = v_1 \frac{S}{N - V_1} - v_2 \frac{VS_1}{V_1}$$
$$\frac{dVS_2}{dt} = v_2 \frac{VS_1}{V_1} - \omega_1 VS_2$$
$$\frac{dWS_1}{dt} = \omega_1 VS_2 - \omega_2 WS_1$$
$$\frac{dWS_2}{dt} = \omega_2 WS_1$$

$$\frac{dR}{dt} = -\Omega_1 R$$
$$\frac{dWR_1}{dt} = \Omega_1 R - \Omega_2 WR_1$$
$$\frac{dWR_2}{dt} = \Omega_2 WR_1$$

$$\frac{dV_1}{dt} = v_1$$
$$\frac{dV_2}{dt} = v_2$$

For those in the classes where the vaccines generate protection (VS1, VS2 and WS1), the degree of protection is determined by the ratio of AstraZeneca (ChAdOx1) vaccine to mRNA vaccines (either Pfizer BNT162b2 or the Moderna COVID-19 vaccine) that has been given to that age group (see Table 1). If a vaccinated individual becomes infected, their probability of being admitted to hospital or dying - which normally only depends on age - is modified by the appropriate vaccine efficacy according to the ratio of the two vaccine types. Booster vaccinations are implemented by moving individuals from the vaccinated or waned class into the recovered class where the level of protection is enhanced. Waning from the booster state is assumed to occur at a low rate comparable to that of recovered individuals(S2).

#### Parameter Inference

Key to the accuracy of any model are the parameters that underpin the dynamics. With a model of this complexity, a large number of parameters are required. Some, such as vaccine efficacy, are assumed values based on the current literature; while others, such as the level of precautionary behaviour over time, are inferred from the epidemic dynamics. Bayesian inference, using an MCMC process, is applied to each of the seven NHS regions in England to determine posterior distributions for each of the regional parameters (further details are given in [14]). The distribution of parameters leads to uncertainty in model projections, which is represented by the 95% prediction interval in all graphs (this interval contains 95% of all predictions).

As the epidemic has progressed, new posterior distributions based on the latest data are initialised from previous MCMC chains – ensuring a rapid fit to historical data. We match to six observations: hospital admissions, hospital occupancy, ICU occupancy, deaths, proportion of pillar 2 (community) test that are position, and the proportion of pillar 2 tests that are S-gene positive (as a signal of the ratio of wild-type to Alpha variant, then a signal of the ratio of Delta to Alpha variant, and more recently a signal of Omicron to Delta). Although not part of the transmission dynamics, these six quantities for each

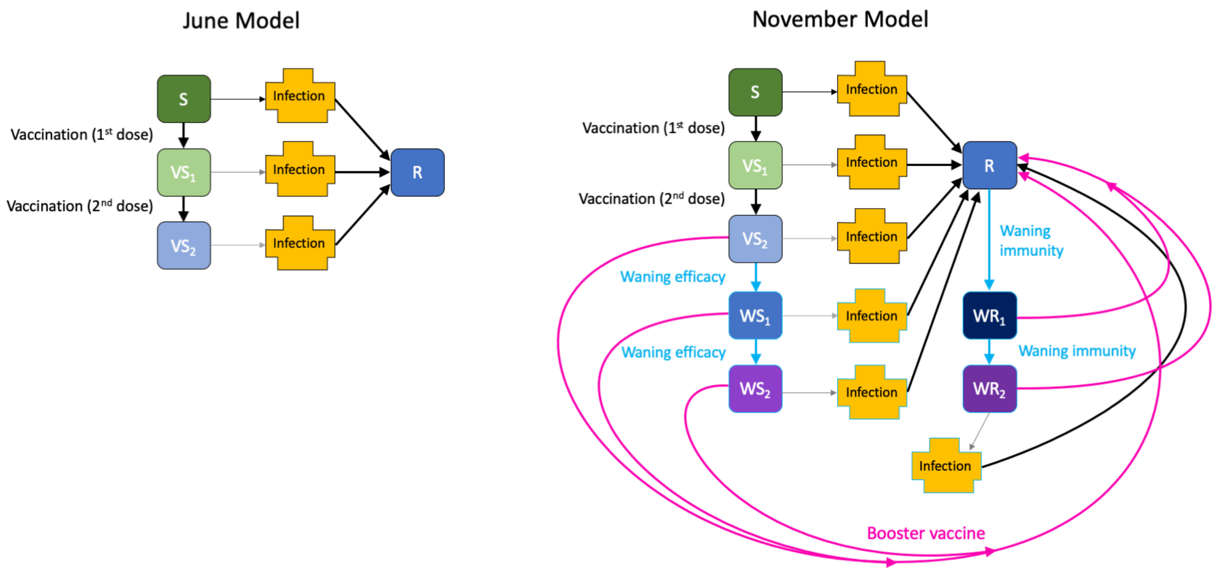


**Fig. S2: Graphical representation of the two models used.** The June model (left) has four non-infected classes: Susceptible, Vaccinated with one dose, Vaccinated with two doses and Recovered; the November model (right) has increased complexity by adding waning of immunity (light-blue) and booster vaccination (pink). For both models there is the added dimension of different variants and more compartments within the infection class.

region can be generated from the number, age and type of infection within the model. Observations and model results are compared by considering the likelihood of generating the observations assuming they are Poisson distributed with a mean given by the results of the deterministic model.
